# Does later chronotype cause poorer adolescent mental health? An Adolescent Brain Cognitive Development (ABCD) Study

**DOI:** 10.1101/2025.10.08.25337391

**Authors:** Rachel Visontay, Hollie R. Byrne, Emma K. Devine, Mirim Shin, Emiliana Tonini, Gabrielle Hindmarsh, Joanne S. Carpenter, Ty Brumback, Lindsay M. Squeglia, Louise Mewton, Ian B. Hickie, Jacob J. Crouse

## Abstract

**Objective:** This study investigated whether chronotype (biobehavioral preference for sleep and wake timing) across early adolescence impacts mental health symptoms at age 13-14. This is a relationship with growing correlational evidence but limited causal exploration.

**Methods:** Participants were 7,489 adolescents (aged 9-10 at baseline; 13-14 at the fifth assessment wave) from the Adolescent Brain Cognitive Development Study (ABCD). Marginal structural models with machine learning-based weight estimation were used to assess the causal impact of chronotype at ages 11-12 and 12-13, as measured by the youth-reported Munich Chronotype Questionnaire, on several dimensions of mental health symptoms at age 13-14 (parent-reported Child Behavioral Checklist internalizing and externalizing values; child-reported Prodromal Psychosis Scale).

**Results:** There was no effect of chronotype at age 11-12 on any outcome. However, later chronotype at age 12-13 was associated with more severe externalizing symptoms (b=.31, p=.02) and prodromal psychosis symptoms at age 13-14 (b=.06, p<.01), but not with more severe internalizing symptoms. Post-hoc analyses indicated the lack of relationship with internalizing held for both anxiety and depression symptoms and in both sexes.

**Conclusions:** There are likely causal effects of adolescent chronotype on mental health symptoms, but these are dependent on the dimension of mental health and period of adolescence. The typically-reported association between later chronotype and more severe internalizing symptoms may not manifest until later adolescence (such as with the common post-pubertal shift in chronotype).

## INTRODUCTION

Rates of adolescent mental illness in high-income countries have dramatically increased in recent years. In the U.S., the prevalence of mental disorders in 10–19-year-olds rose from 16,373 cases per 100,000 people in 2009, to 21,016 per 100,000 in 2021 – an increase of 28% (1). Globally, it is estimated that mental illness accounts for 15% of disease burden in that same age group (2). Given these trends, The Lancet Psychiatry Commission on Youth Mental Health has emphasized that identification of risk factors for mental illness in young people is urgent (3).

One potentially important but understudied factor is the role of chronotype – that is, one’s biobehavioral preference for timing of sleep, wakefulness, and activity (4). Epidemiological studies report associations between chronotype and mental health in a range of populations, particularly between a later chronotype (characterizing so-called ‘night owls’) and poorer mental health (5). In adolescence specifically, later chronotype has been linked to both internalizing symptoms (e.g., mood, anxiety) and externalizing symptoms (e.g., substance use, conduct problems) (6–10). Such relationships generally persist even when accounting for sleep duration (7–9), suggesting there may be something about later sleep-wake timing preference itself – independent of its effects on sleep lag/debt during the school week – that has negative consequences for mental health. Possible mechanisms have been theorized, including greater opportunity for rumination, increased negative thought content, lack of available social support at night, and disturbed circadian rhythms (misalignment of the internal body clock with the external environment) that may cause hormonal changes such as higher morning cortisol (11,12).

However, these cross-sectional studies fail to isolate the direction of effect; longitudinal data are a minimum first step in gathering evidence for a *causal* effect of chronotype on mental health. While some longitudinal studies have been conducted, most have featured small samples and broad age ranges, reporting mixed findings. For example, some studies have found that later chronotype predicts subsequent depressive symptoms in adolescents (13,14), while others have found it does not (15), or that such effects do not survive when accounting for baseline symptoms (16). Strengthening causal inference in this research area calls for longitudinal data with large samples and the ability to isolate effects at specific ages, to avoid masking heterogeneity across the adolescent period.

Importantly, identifying causal relationships using improved statistical methods, such as marginal structural models (MSMs), is also needed. MSMs attempt to fully adjust for measured confounders by assigning weights to each participant prior to regression modelling to balance confounders across exposure levels (i.e., different chronotypes). These weights are known as inverse probability of treatment weights (IPTWs). In our example, IPTWs capture the probability of an individual having their reported chronotype given their values on relevant covariates (e.g., age, sex). The less typical an individual is for their particular chronotype, the greater the weight they are assigned. When these weights are applied to the sample, a ‘pseudo population’ is created wherein the covariate distribution across chronotypes is balanced (17).

Exploring the chronotype–mental health relationship in adolescence is particularly challenging given that these factors are in flux: puberty brings changes to both biological factors (e.g., delayed melatonin release, heightened circadian-sensitivity to light) and environmental factors (e.g., social interactions, screen use) that trigger the delay of sleep timing (11), while also being the peak period of onset for mental illness (18). MSMs are well-equipped to deal with such dynamic developmental periods where exposures and outcomes fluctuate, and certain factors (e.g., sleep disturbances, screen use) act as both relationship confounders and mediators over time. For example, with reference to the Directed Acyclic Graph in Figure 1, screen use at age 11-12 may confound the relationship between age 12-13 chronotype and age 13-14 mental health symptoms, while also itself being affected by age 11-12 chronotype. Traditional regression adjustment approaches that control for screen use at age 11-12 would bias results by adjusting away the effect of age 11-12 chronotype mediated through screen use, while a lack of control for screen use at age 11-12 would lead to results biased by confounding. By contrast, the weighted MSM, in rebalancing covariates but not fixing their values, avoids this problem.

**Figure 1.**
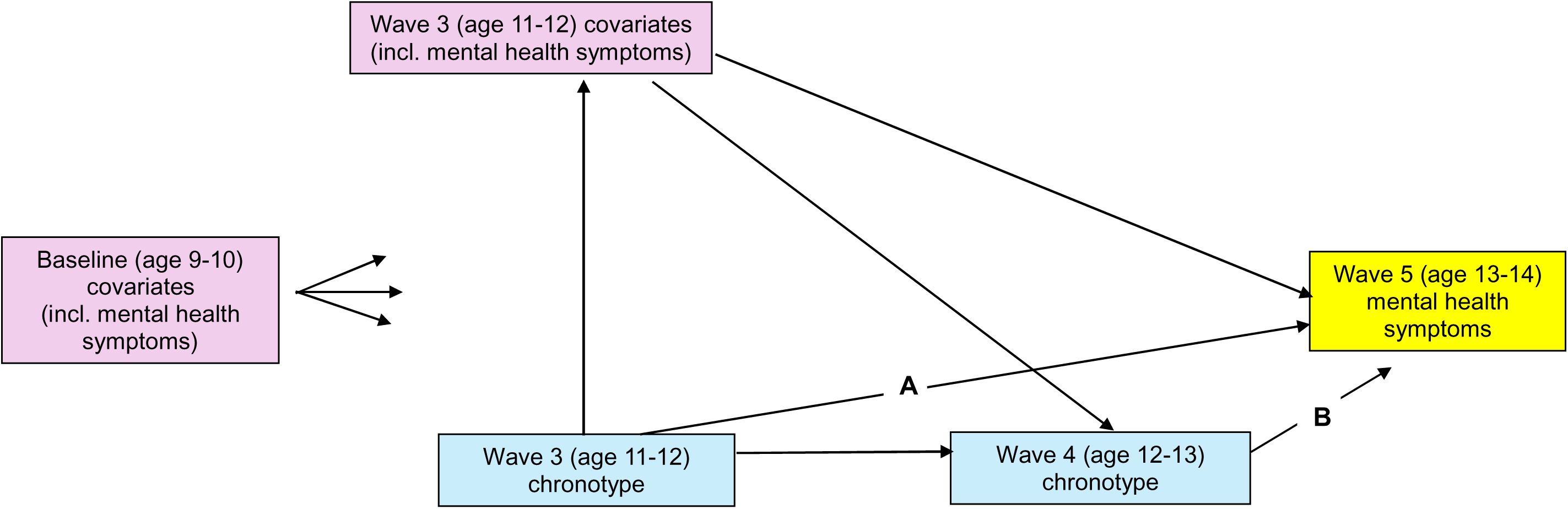
Directed acyclic graph depicting theorised relationships between variables over time^a^. ^a^The MSM approach models the causal pathways A and B, using weights to control for the influence of the other depicted pathways.

The aim of this study was to apply an MSM approach to evaluate the impact of chronotype at age 11-12 (when chronotype was first measured) and age 12-13 on mental health symptoms at age 13-14 in a very large, developmentally-sensitive cohort (the Adolescent Brain Cognitive Development Study; ABCD (19)). It was hypothesized that later chronotype at each time point would be a potentially causal factor for poorer mental health across dimensions (internalizing, externalizing, prodromal psychosis) at age 13-14. Analyses and hypotheses were pre-registered: https://osf.io/2ndvp.

## METHODS

### Study Design and Participants

ABCD is a multi-site (N=21) longitudinal study following the neurodevelopment, health, and lifestyles of a large adolescent cohort in the U.S. Children were recruited when aged 9-10-years, from 2016-2018. The ABCD 5.1 data release was used, which contains complete data up to the fourth assessment wave (N=11,868; age 12-13) and a subset of data for the fifth assessment wave (N=4,754; age 13-14). Parents or caregivers provided informed consent and all participants gave assent. The ABCD protocol was approved by the centralised institutional review board (IRB) at the University of California, San Diego and by the IRBs at each of the 21 sites.

For the present study, siblings were removed at random such that only one participant per family remained (see Figure 2). From there, eligible individuals were those who had valid baseline covariate data (including pre-existing values of mental health outcomes), and chronotype data at age 11-12. The MSM incorporated chronotype at age 11-12 and age 12-13, covariates and mental health values at baseline and age 11-12, and the final mental health outcomes at age 13-14. In addition to those who dropped out of the study completely, participants missing chronotype data at age 12-13, covariate data at age 11-12, or outcome data at age 13-14 were considered censored at those respective time points. This left a final sample of 2,668/2,698 (depending on outcome assessed).

**Figure 2.**
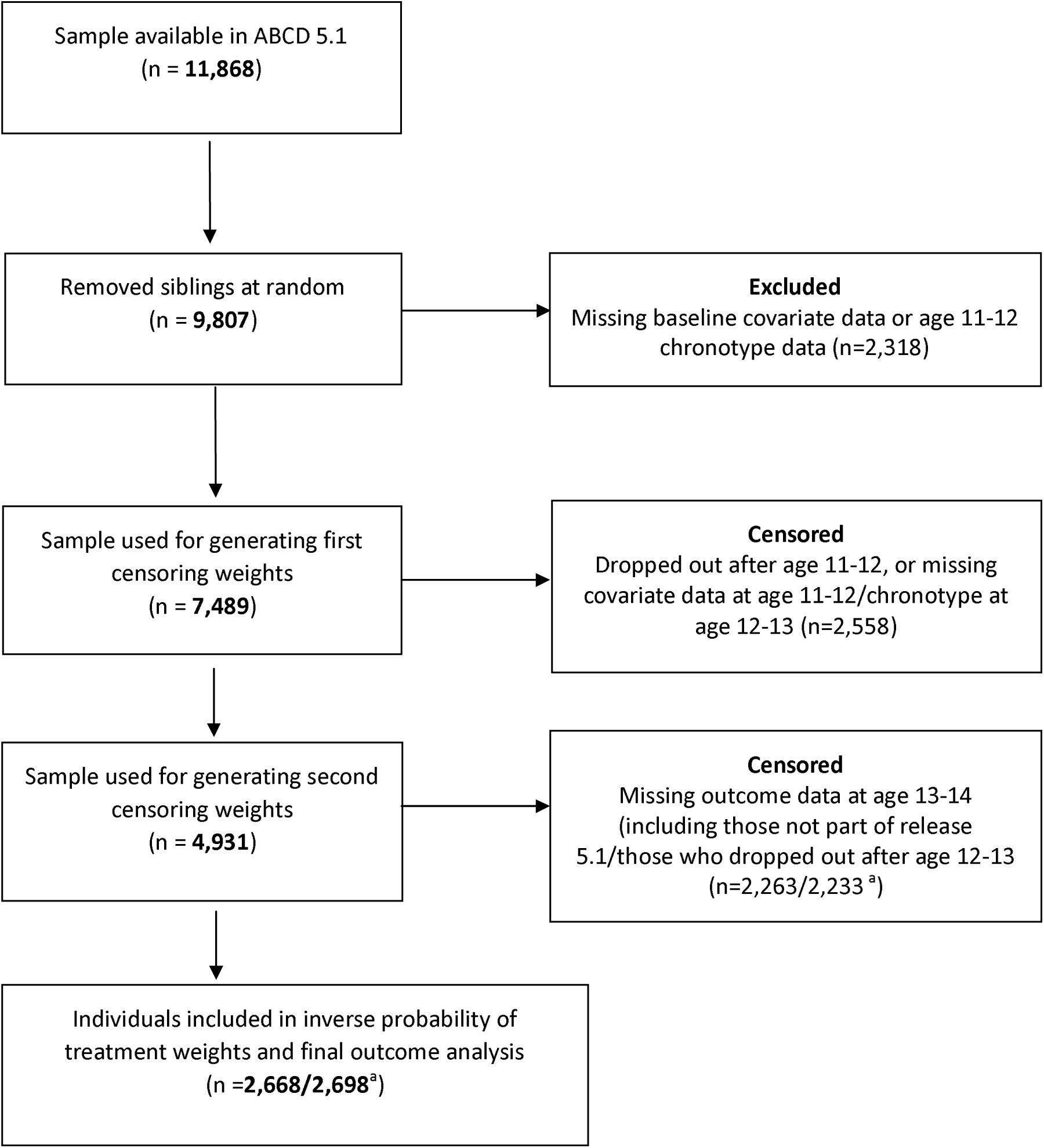
Eligibility flow chart. ^a^First number applies to CBCL outcomes analysis; second to psychosis outcome analysis

### Measures

#### Mental health outcomes

Three outcome variables were included, reflecting different domains of mental health. These were the Child Behavior Checklist’s (CBCL) (20) internalizing and externalizing T-scores (the sum of parents’ responses about their child to all internalizing questions/externalizing questions of the CBCL, standardized according to the age and sex of the child), and the sum score on the youth-reported Prodromal Psychosis Scale (21) (PPS; log-transformed to account for skewness). The CBCL internalizing sum score combines syndrome scales that capture anxious-depressed, withdrawn-depressed, and somatic complaints symptoms, while the externalizing sum score combines syndrome scales that capture rule-breaking and aggressive behavior. Each outcome was used in its continuous form.

#### Chronotype

Chronotype was assessed using the youth-reported Munich Chronotype Questionnaire (MCTQ) (22), with the derived variable ranging from 16-40 (higher scores represent later chronotypes). Using the MCTQ, chronotype is based on actual sleep timing on free days, which seems to be a stronger predictor of mental health symptoms than self-reported ‘preferences’ (23). See the online supplement for details of chronotype score generation.

#### Covariates

Fixed baseline covariates included child’s age, sex, and level of parental education. Time-varying covariates assessed at baseline and age 11-12 were body mass index (BMI), parent-reported child pubertal development (24), parent-reported child sleep disturbances (25), youth-reported average sleep duration, youth-reported average daily screen time, as well as preceding values of the mental health outcomes. More detail on these covariates and their derivation is available in the online supplement. As individuals were clustered within 21 recruitment locations, study site was also included in analyses.

### Statistical Analyses

#### Marginal structural models

MSMs involved the sequential generation of IPTWs for chronotype at age 11-12 and chronotype at age 12-13. Given the likelihood of differential attrition across follow-up (i.e., those with poorer mental health outcomes being more likely to drop-out of ABCD), inverse probability of censoring weights (IPCWs) were also calculated for censoring status between age 11-12 and age 12-13, and censoring between age 12-13 and age 13-14. IPCWs allow the final MSM to be representative of the initial sample. For each individual, an overall weight was generated by multiplying all IPTWs and IPCWs together. The final analysis involved a standard outcome regression weighted by these overall weights, *without* additional covariate adjustment.

#### Weight generation

IPTWs and IPCWs were generated using all relevant covariates from previous waves (covariates measured at the same timepoint as exposure may be consequences, rather than confounders, of chronotype). Machine learning-based methods for weight estimation were used, meaning that the functional form of relationships between covariates and chronotype were determined in a data-driven manner (avoiding the possible misspecification that occurs when the researcher explicitly nominates a functional form in traditional weight estimation). Given machine learning models cannot accommodate random factors, study site was entered as a fixed effect, and all interaction terms monitored for adequate balance – an established method for incorporating clustering into propensity score generation (26). The weights were trimmed at the 95^th^ percentile to mitigate the impact of very large ones, and were calculated separately for each of the three mental health outcomes. IPTWs greatly improved balance across chronotypes, although the conservative 0.1 correlation rule of thumb was not achieved for all covariates (particularly for their interactions and polynomials). Censored individuals contributed to the generation of IPCWs but were removed prior to the final outcome analysis. IPCWs achieved balance between censored and uncensored groups for all covariates within a standardized mean difference of 0.1. See the online supplement for further information on weight generation and balance achieved after weighting.

#### Linear regression

Final weights were applied to multivariable linear regression models where the mental health outcome was regressed on chronotype measured at age 11-12 and age 12-13. Separate models were run for each of the mental health outcomes, using the relevant weights. Cluster-robust standard errors were calculated to account for both the variability introduced by weights and the clustered natured of the data (sufficient given low intraclass correlation coefficients; see Table S4).

All analyses were conducted in R, version 4.4.2. The *WeightIt* package (27) was used for calculating IPCWs and IPTWs, and the *survey* package (28) for running the weighted regression (MSM) models. Code is available here: https://github.com/Hi-Vis/Chronotype-and-mental-health.

#### Deviations from pre-registration

The analyses here deviated from the pre-registered protocol in a few ways. Firstly, some of the pre-registered covariates were not included in final IPTWs. Childhood trauma variables were not available at baseline. Brain-based structural MRI features displayed very low imbalance across chronotypes and substance use at baseline/age 11-12 was very minimal, thus both sets of variables were removed (a smaller covariate pool is more conducive to achieving balance). We had also originally proposed Kiddie Schedule for Affective Disorders and Schizophrenia (K-SADS) categorical diagnoses as additional outcomes, but anomalies in the 5.1 ABCD release meant they could not be included. Finally, the cumulative effect of chronotype across ages 11-12 and 12-13, beyond the separate effects of chronotype at each of the two waves, was not calculated due to differing effect directions at the two waves.

## RESULTS

### Study sample

At baseline, the sample (n=11,868) had a mean age of 9.92 years (SD=0.62) and 45.9% were female. Sample characteristics for values of key time-fixed and time-varying covariates at baseline are presented in Table 1. In the final sample used for the substantive analysis (n=2,668), the mean chronotype at age 11-12 was 27.58 (SD=1.66), i.e., a sleep midpoint of 3.30am, and at age 12-13 was 28.03 (SD=1.94), i.e., a sleep midpoint of 4am.

**Table 1.** Selected baseline characteristics of eligible individuals in a nationally representative cohort of US youth.

| Characteristic | Uncensored final sample<sup>a</sup> |  | Full baseline sample |  |
| --- | --- | --- | --- | --- |
|  | (N=2,688) |  | (N=7,489) |  |
|  | Mean | SD | Mean | SD |
| Age | 9.92 | 0.61 | 9.92 | 0.62 |
| BMI | 18.6 | 3.88 | 18.8 | 4.07 |
| Pubertal Development Scale and<br>Menstrual Cycle Survey History<sup>b</sup> | 1.68 | 0.84 | 1.74 | 0.86 |
| CBCL Internalizing T-Score | 48.6 | 10.3 | 48.8 | 10.6 |
| CBCL Externalizing T-Score | 45.3 | 9.82 | 45.7 | 10.2 |
| Prodromal Psychosis Severity Score | 5.78 | 10.0 | 5.96 | 10.2 |
|  | N | % | N | % |
| Sex (Female) | 1204 | 45.1 | 3475 | 46.4 |
| Parental Education |  |  |  |  |
| Less than HS degree/GED equivalent | 118 | 4.4 | 394 | 5.3 |
| HS degree/GED equivalent | 199 | 7.5 | 710 | 9.5 |
| Some college/Associate's degree | 717 | 26.9 | 2117 | 28.3 |
| Bachelor's degree | 856 | 32.1 | 2185 | 29.2 |
| Master's degree | 580 | 21.7 | 1556 | 20.8 |
| Professional school or doctoral degree | 198 | 7.4 | 527 | 7.0 |

<sup>a</sup> Based on the sample used for the CBCL outcomes
<sup>b</sup> Higher scores indicate more advanced pubertal development

### MSM results

The results of the three MSM models are displayed in Table 2. For internalizing symptoms, neither age 11-12 chronotype nor age 12-13 chronotype had a significant effect on symptoms at age 13-14, with age 11-12 chronotype displaying a non-significant *negative* effect on symptoms at age 13-14 (later chronotype being protective; b=-0.174; *p*=0.362). By contrast, for both externalizing and prodromal psychosis symptoms, age 12-13 chronotype was significantly associated with increased symptoms at age 13-14 (later chronotype *causing* increased symptoms; b=0.312; *p=*0.020, b=0.055; *p*=0.002, respectively). Age 11-12 chronotype was not significantly associated with either externalizing or prodromal psychosis symptoms.

**Table 2.** Results of marginal structural model analyses regressing mental health outcomes at age 13-14 on chronotype in early adolescence.

|  | **Estimate**<br>**(unstandardized)** | **Robust std. error** | **P value** |
| --- | --- | --- | --- |
| **Internalizing** |  |  |  |
| Age 11-12 chronotype | -0.17397 | 0.18611 | 0.362 |
| Age 12-13 chronotype | 0.07782 | 0.16946 | 0.651 |
| **Externalizing** |  |  |  |
| Age 11-12 chronotype | -0.1116 | 0.1813 | 0.5454 |
| Age 12-13 chronotype | 0.3116 | 0.1222 | 0.0196 |
| **Prodromal Psychosis** |  |  |  |
| Age 11-12 chronotype | 0.02492 | 0.01328 | 0.07598 |
| Age 12-13 chronotype | 0.05506 | 0.01534 | 0.00195 |

### Post-hoc analyses

Given the unexpected lack of relationship between chronotype and internalizing symptoms, post-hoc analyses were conducted in which 1) the CBCL internalizing T-score was replaced with the DSM-5 oriented depression and anxiety sum T-scores (given some evidence of differing associations with chronotype between the two constructs (29)), and 2) the sample was split by sex (weights were regenerated given the different sample) to explore whether associations differed by sex. There were no significant relationships for either subscale (Table S1). When the sample was split by sex, there was still no significant relationship with internalizing in either sex (Tables S2 and S3), with the only remaining significant relationship being for prodromal psychosis in males. Finally, a correlation between externalizing and prodromal psychosis symptoms was conducted to ascertain the extent of overlap in these populations (r=.09, p<.001),

## DISCUSSION

In a large developmentally-informative cohort of adolescents from the general population, this causal inference analysis of the relationship between chronotype in early adolescence and mental health symptoms at age 13-14 found small, but likely causal, effects of later chronotype at age 12-13 on externalizing and prodromal psychosis symptoms, independent of sleep duration and sleep problems. No effects were found for internalizing symptoms, nor of chronotype at age 11-12 on any mental health outcome. This remained true in post-hoc analyses using anxiety and depression-specific subscales of internalizing, and when stratifying by sex.

The effects of chronotype on externalizing symptoms are consistent with the associational literature described earlier. The size of the effect is small but meaningful (although it should be noted that significance was not maintained in the smaller, sex-specific samples). One potential interpretation is the greater opportunity for misbehaviour (getting into trouble) in the evening (30). Other key hypothesized mechanisms include poorer adjustment and increased conflict because of circadian disturbances and misalignment of biological and social rhythms, or the effects of circadian disturbance on cortisol and serotonin release, as well as the serotonergic system (31). Also possible is that the two phenomena (chronotype and externalizing symptoms) have common causes, i.e., shared personality factors such as higher risk-taking and sensation seeking, or shared genetic factors (32).

While the association found between later chronotype and prodromal psychosis is also consistent with much of the literature in young people (33), our findings confirm the directionality of the effect in a causal framework. One hypothesis underlying this finding is that circadian disturbances may interact with genetic risk and environmental stressors to contribute to psychosis risk, mediated by changes to cortisol response or neurodevelopment (34). The small correlation between scores on the externalizing and prodromal psychosis scales suggests two separate populations contributing independently to the chronotype–psychosis and chronotype–externalizing effects.

The lack of observed relationship between later chronotype and more severe internalizing symptoms was surprising. It may mean this oft-reported relationship is not causal, although most of the hypothesized mechanisms proposed for the observed increases in externalizing and psychosis symptoms (e.g., cortisol levels) should theoretically apply to internalizing symptoms too. More likely is that the association exists but may not manifest until later adolescence/adulthood. In support of this, we note that the normative developmental chronotype shift (toward later in the evening) has only just begun by age 12-13, with late chronotype peaking (and therefore greater variation in the exposure of interest) only later in adolescence (35). Indeed, recent research in adults suggests that later chronotype’s effect on psychopathology may be largely mediated by the phenomenon of sleep inertia – difficulty transitioning from sleep to wake (cognitive sluggishness), which is more likely to be experienced by those with later chronotypes forced to wake during their biological night (36). Without having yet reached that substantial shift in chronotype, responsible for a clash between biological night and morning demands, we would not expect as great an effect in this younger (and more morning-like) age group.

Finally, the fact that only chronotype at age 12-13 but not 11-12 influenced externalizing and psychosis symptoms at age 13-14 could indicate that only more proximal sleep-wake timing is important for mental health outcomes. Alternatively, there may be something particular about age 12-13 specifically but not 11-12, occurring. For example, acceleration in chronotype shift from age 13 (37) may mean that there is too little variation in chronotype prior to that for an effect to manifest. Alternatively, the start of more demanding schedules as children enter middle school may lead to a greater misalignment (and greater repercussions of that misalignment) between biological clocks and social timing.

### Strengths

The application of causal inference methods to the relationship between chronotype and mental health is a fledgling undertaking. The present study contributes preliminary evidence that there may be a causal effect of later chronotype on increased symptoms of psychopathology in adolescents. The rich characterization of the ABCD cohort meant that the present study was able to control for a wide range of variables that may confound the chronotype–mental health relationship over time. As opposed to traditional modelling, the use of MSMs appropriately accounts for the dynamic nature of sleep-wake timing and confounding variables in early adolescence.

### Limitations

Despite the rich nature of the ABCD cohort, there may be additional important covariates we did not or could not include. For those variables that were included, while the weights greatly improved balance, they did not always bring covariate imbalance within the .1 standardized mean difference rule of thumb threshold, meaning that confounding may still be biasing results. The CBCL outcome measures are also limited by being parent-reported, which may not accurately reflect experienced symptoms (particularly in the case of internalizing).

Regarding improved causal inference, in our study, the MSM only offers advantages related to longitudinal confounding/mediation (as depicted in Figure 1) if chronotype at age 11-12 affects mental health/covariates at the same age (indeed, it is likely that the relationship between chronotype and mental health symptoms is bidirectional at any given timepoint). While unlikely, if this assumption does not hold, the use of machine learning-estimated IPTWs still offers advantages over traditional regression adjustment in producing a marginal (as opposed to conditional) interpretation, and a reduced risk of misspecifying covariate–exposure relationships.

### Future directions

The present study demonstrates the importance of isolating discrete timepoints within adolescence and discrete domains of psychopathology when examining chronotype and mental health, given heterogenous effects. The recent release of ABCD 6.0, which provides data on later ages in adolescence (up to 15-16) provides an opportunity to reassess the impact of chronotype as shifts in sleep timing become more pronounced, as do increases in internalizing symptoms. Incorporation of these additional waves would exploit the full potential of MSMs to deal with longitudinal complexity.

Replacing parent-reported CBCL outcomes with child-reported K-SADS data using the 6.0 release also merits testing, as the latter may offer clearer clinical significance. Additionally, although self-rated chronotype correlates well with biospecimen-derived markers of circadian timing, such as dim-light melatonin onset (38), looking at this research question using a more objective measure of sleep-wake timing, e.g., from wearables, would be valuable (39). Methodologically, a causal mediation analysis testing the roles of sleep inertia and hormone levels on the path from chronotype from mental health would be of great interest. So too would performing a MSM focused instead on the effect of mental health on chronotype, given likely bidirectionality of the relationship (11,40).

Finally, triangulating findings with analyses using other causal inference methods with complementary strengths and weaknesses, and across cohorts, is required. Of particular promise are Mendelian Randomization analyses (a genetically-informed technique), that so far have only been used to show causal effect of chronotype on mental health in *adult* populations (41–43).

## Conclusion

It appears that there are likely causal effects of adolescent chronotype on mental health symptoms, but that these are dependent on dimension of mental health, and period of adolescence. There is potential for intervening on chronotype to prevent externalizing and psychosis symptoms, as well as for delaying school start times to mitigate the misalignment between circadian timing and social rhythms that potentially underpin the relationship.

## Data Availability

All data are available online by applying to ABCD.

## Disclosures

IBH is Co-Director, Health and Policy, at the Brain and Mind Centre, University of Sydney, Australia, which operates an early-intervention youth services at Camperdown under contract to headspace. He has previously led community-based projects and pharmaceutical industry–supported projects (Wyeth, Eli Lily, Servier, Pfizer, AstraZeneca and Janssen Cilag) focused on the identification and better management of anxiety and depression. He is the chief scientific advisor to and is a 3.2% equity shareholder in InnoWell, which aims to transform mental health services through the use of innovative technologies. All other authors report no financial relationships with commercial interests.

## Acknowledgments

RV was funded for this work by an Engagement Collaborative Grant from the Brain and Mind Centre at the University of Sydney. IBH was supported by a National Health and Medical Research Council (NHMRC) Leadership Fellowship (2016346). JJC was supported by a NHMRC Emerging Leadership Fellowship (2008196). LS is supported by the National Institute of Alcohol Abuse and Alcoholism (K24AA031052). MS is supported by philanthropic donations from families who are affected by mental illness (who would like to be left anonymous).

## Notes

### Author Declarations

The ABCD protocol was approved by the centralised institutional review board (IRB) at the University of California, San Diego and by the IRBs at each of the 21 sites.

