## Supplementary material for "Does later chronotype cause poorer adolescent mental health? An Adolescent Brain Cognitive Development (ABCD) Study": online supplement

1. Chronotype calculation

The continuous chronotype variable was defined as midsleep time on free days, corrected for sleep debt from school days, and derived using the questions in the Munich Chronotype Questionnaire that probe average sleep duration on free days, average sleep duration on weekdays, and midsleep point on free days. For those whose sleep duration on free days was less than school days (unusual), their midsleep point on free days was taken as their chronotype value. For those whose sleep duration on free days is greater, the average weekly sleep duration was subtracted from the sleep duration on free days and divided by two, before being subtracted from midsleep point on free days. The resulting chronotype was a score from 0-24. Finally, scores under 16 had 24 added, such that the final range of possible scores was 16-40.

A score of 16 therefore represents a very early chronotype (4pm midpoint of sleep – for someone with an 8-hour sleep duration that would be a 12pm bedtime and 8pm wake-up). A score of 22 represents a 10pm midpoint –a 6pm bedtime and 2am wakeup). A score of 30 represents a 6am midpoint the following day (2am bedtime, 10am wake up). Those who are woken by parents/alarms on free days were considered missing for chronotype value.

2. Derivation of covariates

| COVARIATE | NOTES |
| --- | --- |
| Level of parental education | **demo_prnt_ed_v2** collapsed into 6 categories (less than high school/GED equivalent, some college/associates degree, bachelors degree, masters degree, professional school or postdoctoral degree) |
| BMI | Calculated as **anthroweightcalc**/(**anthroheightcalc**^2)*703 |
| Pubertal development | Pubertal Development Scale and Menstrual Cycle Survey History: development score derived from three sex-neutral questions on height changes, skin changes, and body hair, as well as two sex-specific questions on breast development and menstruation/deepening voice and facial hair. Derived pubertal development score based on parent-reported **pds_1_p**, **pds_2_p**, **pds_3_p**, **pds_f4_p**, **pds_f5b_p**, **pds_m4_p**, **pds_m5_p**, with 1=prepubertal, 2=early, 3=mid, 4=late, 5=post |
| Sleep disturbances | Parent-reported Sleep Disturbance Scale: a binary presence of sleep disturbances was determined by summing individual components related to disorders of initiating and maintaining sleep, sleep breathing, arousal, sleep-wake transition, excessive somnolence, and sleep hyperhidrosis (**sds_p_ss_dims**, **sds_p_ss_sbd**, **sds_p_ss_da**, **sds_p_ss_swtd**, **sds_p_ss_does**, **sds_p_ss_shy** >=39) |
| Average sleep duration | **mctq_sdweek_calc** |
| Average daily screen use | Calculated from self-reported average weekday and weekend use **stq_y_ss_weekday**, **stq_y_ss_weekend**, **screentime_wkdy_typical_hr**,  **screentime_wkdy_typical_min**,  **screentime_wknd_typical_hr**,  **screentime_wknd_t_min** |
| Earlier CBCL and Prodromal Psychosis Scale values | As described in main paper for these scales when used as outcomes |

3. Variables included in IPCWs and IPTWs

For the first IPCW, covariates included baseline age, parental education, sex, and baseline values of average daily screen time (log-transformed), BMI (log-transformed), pubertal development, externalizing t score, internalizing t score, prodromal psychosis symptoms t score (log-transformed), age 11-12 chronotype, and study site. For the second IPCW, covariates included these covariates as well as age 11-12 values of average daily screen time (log-transformed), BMI (log-transformed), pubertal development, externalizing t score, internalizing t score, prodromal psychosis symptoms t score (log-transformed), and age 12-13 chronotype.

As depicted in Figure 1, because there is an assumed causal effect of age 11-12 chronotype on age 11-12 covariates (including mental health symptoms), the latter were not included in IPTW generation for the former. Instead, age 11-12 IPTWs were constructed using baseline covariates (age, parental education, sex, average daily screen time (log-transformed), BMI (log-transformed), pubertal development, externalizing t score, internalizing t score, prodromal psychosis symptoms t score (log-transformed), and study site. Age 12-13 IPTWs were constructed using these covariates as well as age 11-12 values of average daily screen time (log-transformed), BMI (log-transformed), pubertal development, externalizing t score, internalizing t score, prodromal psychosis symptoms t score (log-transformed), and age 11-13 chronotype.

4. Detail on machine learning estimation of weights

SuperLearner was used to estimate IPCWs, using xgboost and glm as base algorithms. Generalised boosted models were used to estimate IPTWs. Balance before and after weighting was assessed for all covariates, as well as their squared values, and all possible interactions.

Figure S1. Covariate balance achieved after IPCW weighting^a^

a) CBCL internalizing – censored between age 11-12 and age 12-13

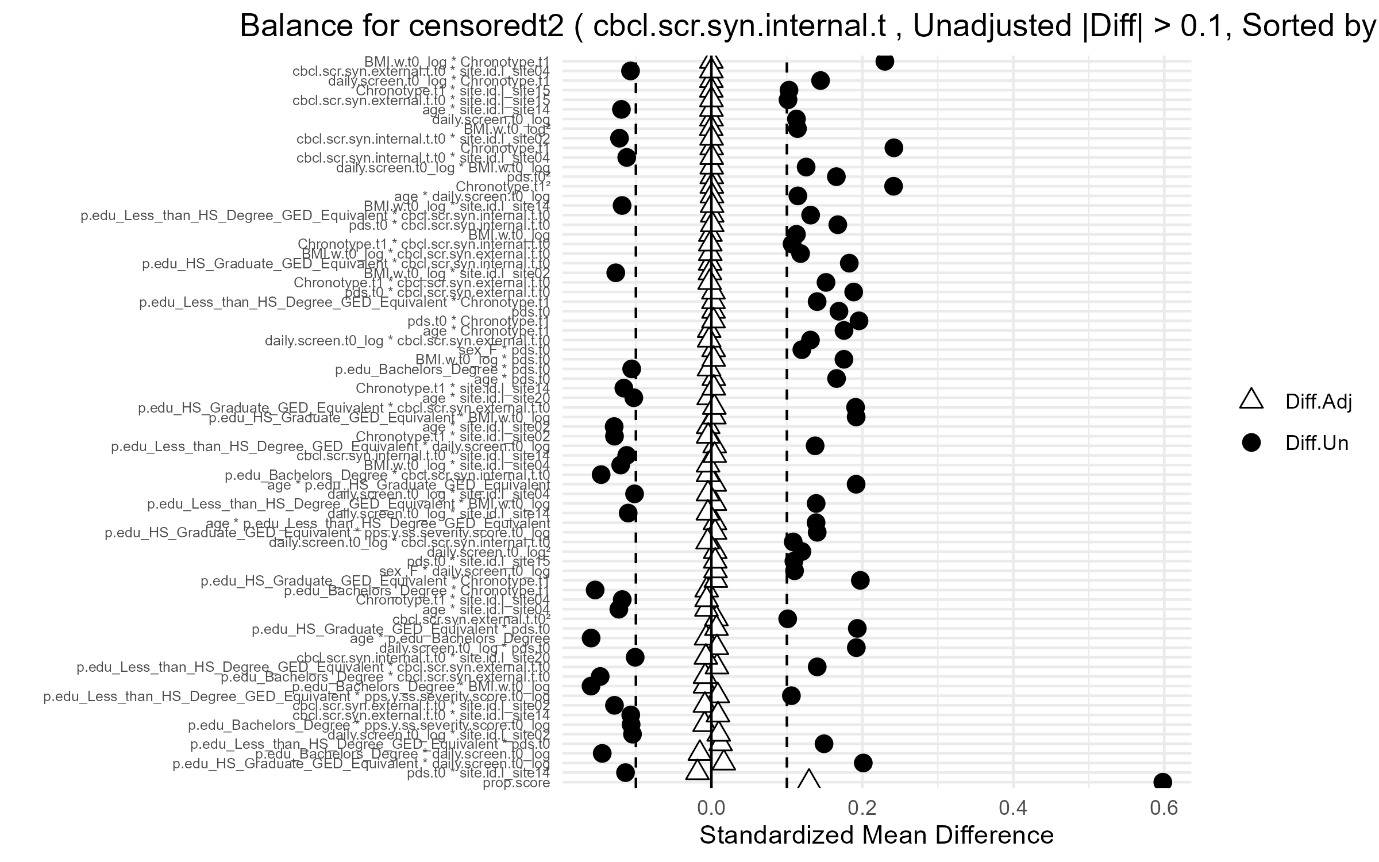

b) CBCL internalizing – censored between age 12-13 and age 13-14

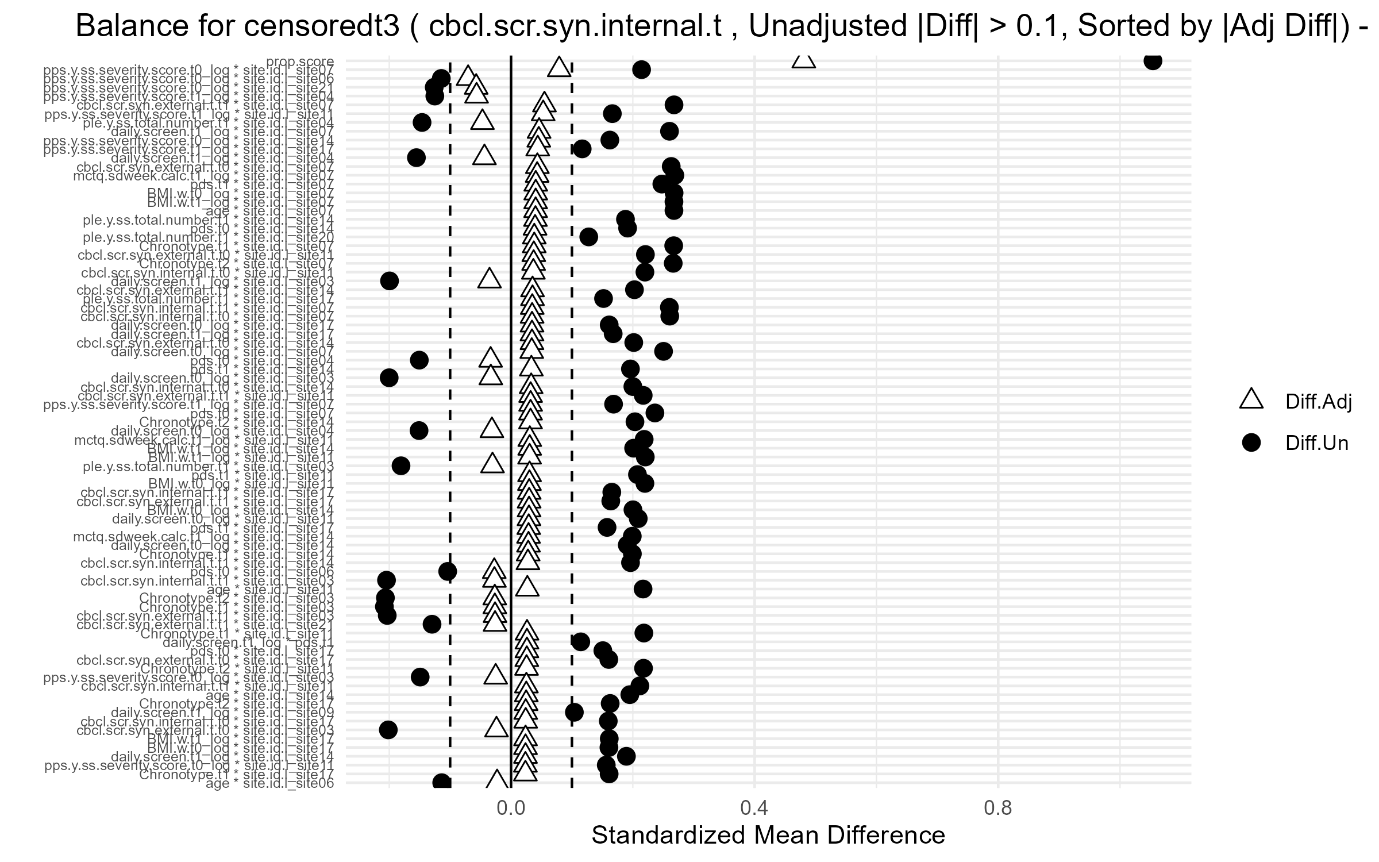

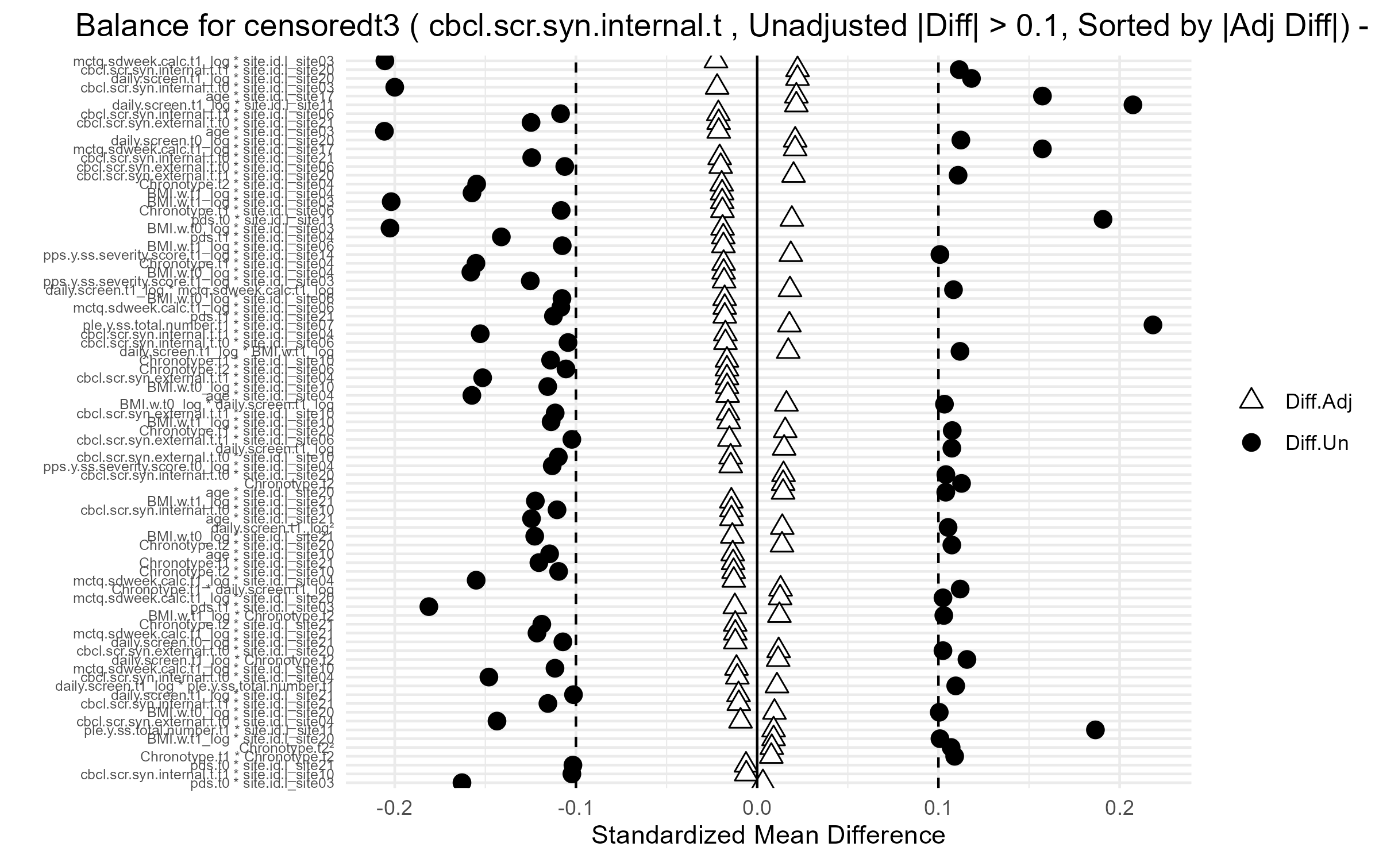

^a^Plots displayed for internalizing analyses only as those for externalizing and prodromal psychosis were almost identical. For simplicity, the above figures display only those covariates (or their interactions/polynomials) that were unbalanced (beyond a 0.1 standardized mean difference) between censored and uncensored individuals prior to weighting.

Figure S2. Covariate balance achieved after IPTW weighting^a^

a) CBCL internalizing – age 11-12 chronotype

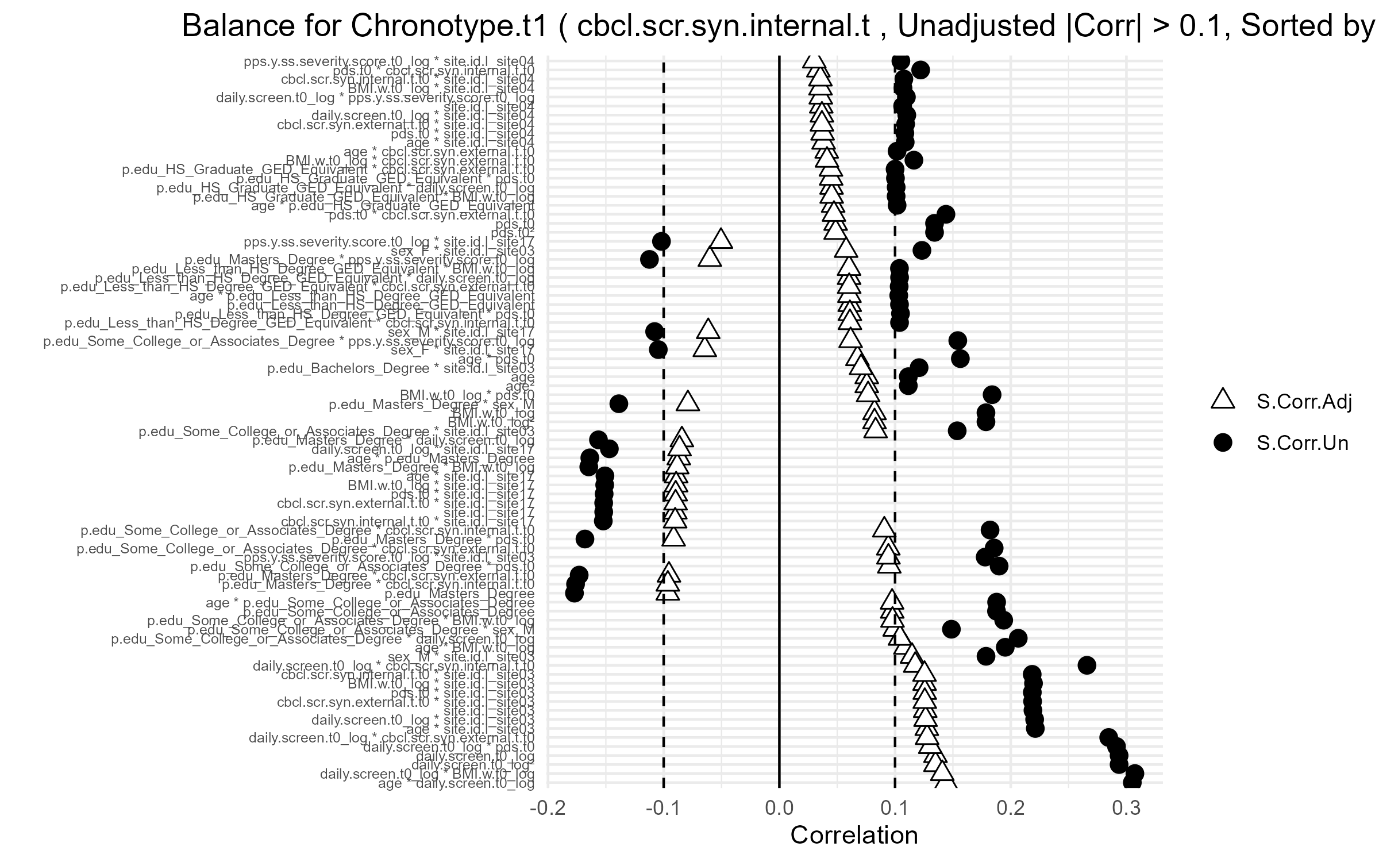

b) CBCL internalizing – age 12-13 chronotype

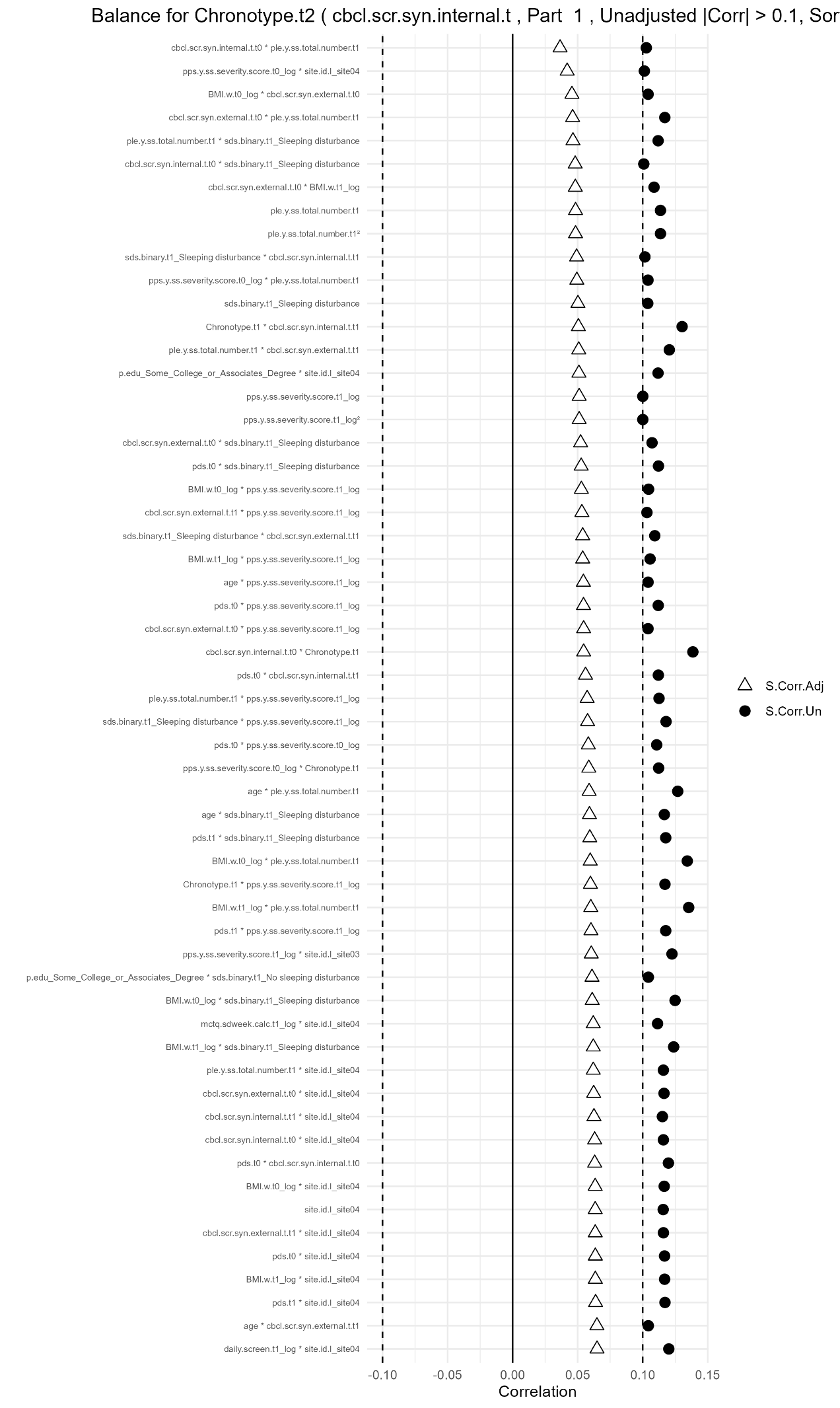

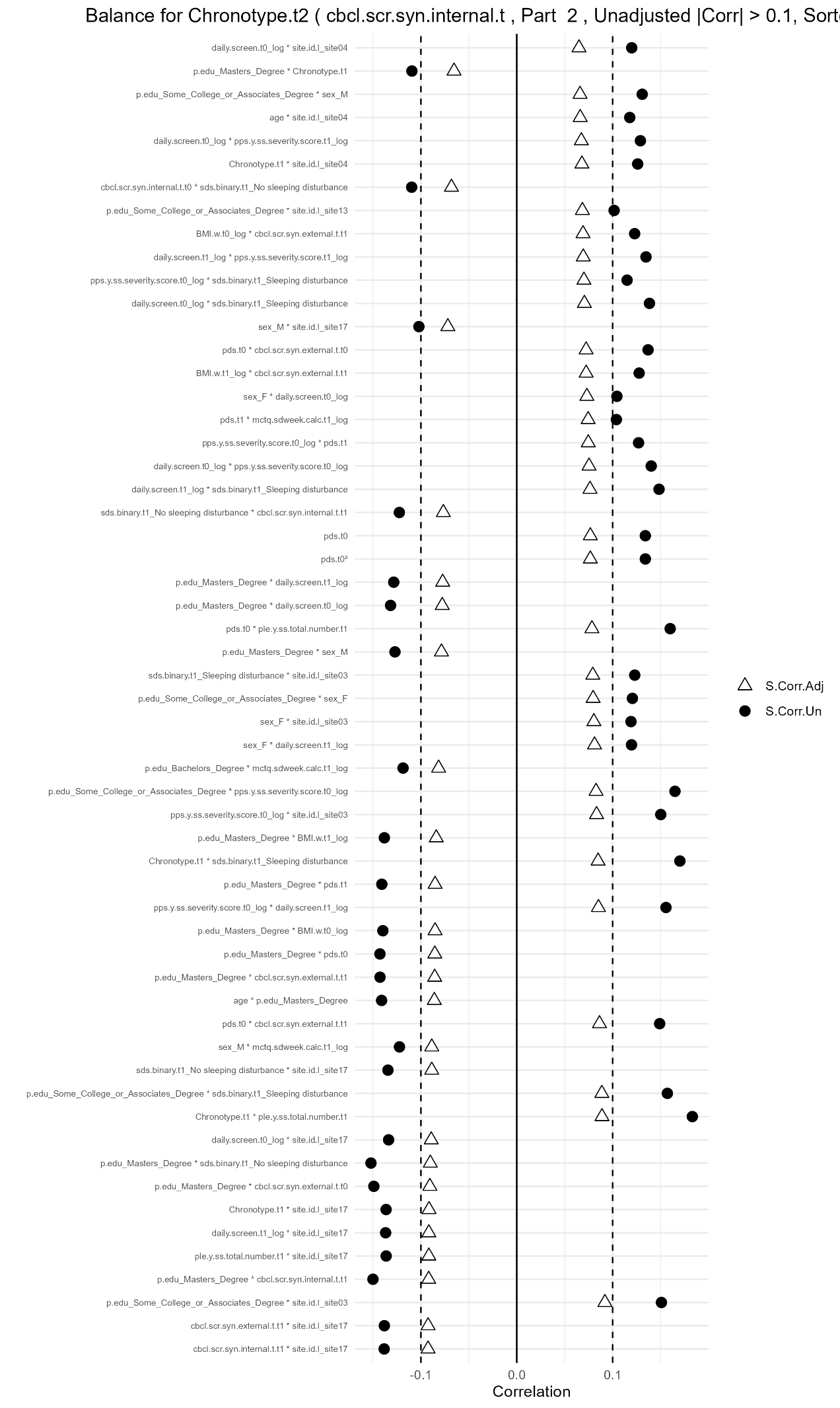

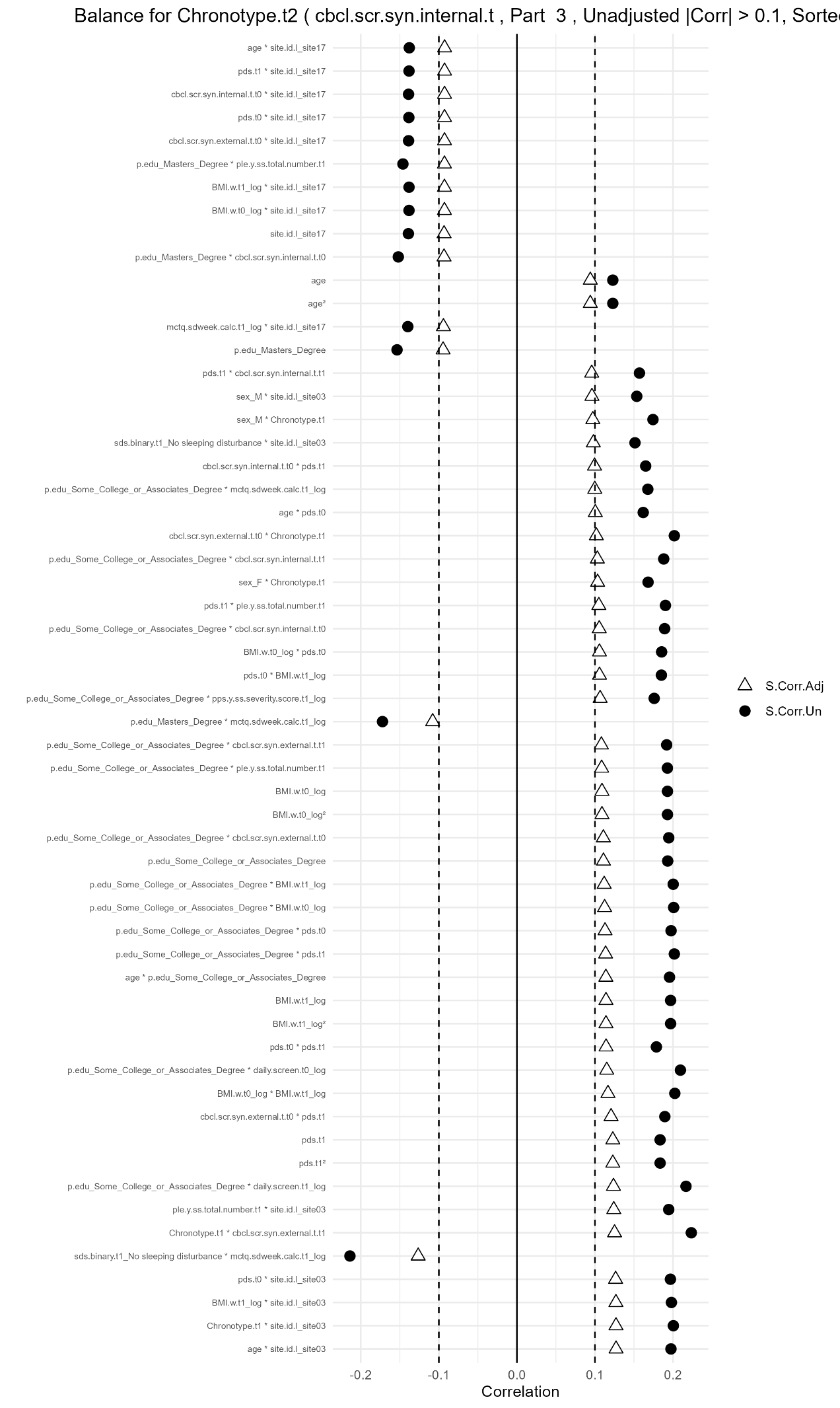

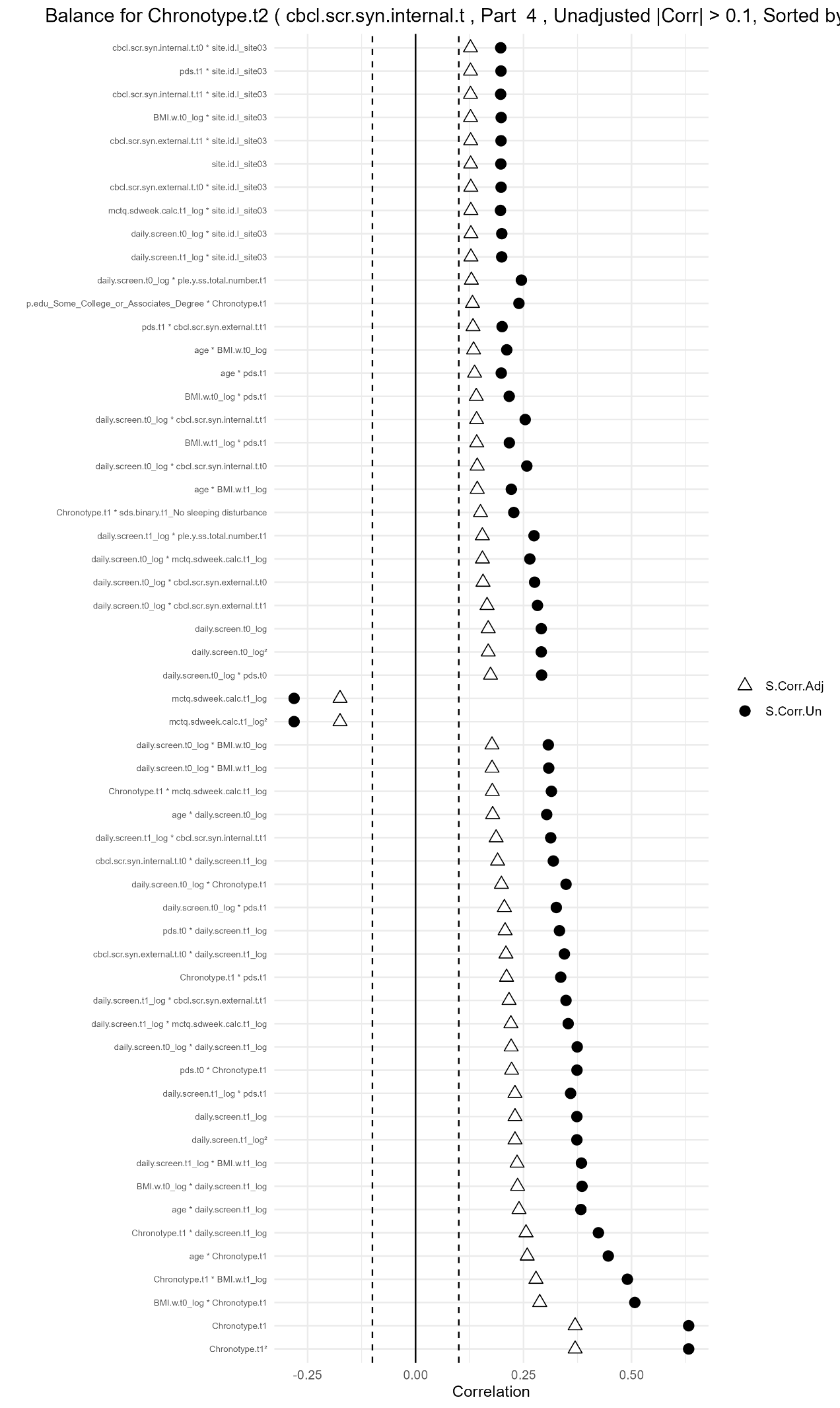

^a^Plots displayed for internalizing analyses only as those for externalizing and prodromal psychosis were almost identical. For simplicity, the above figures display only those covariates (or their interactions/polynomials) that were unbalanced (beyond an |0.1| correlation) across chronotype prior to weighting.

Table S1. Results of post-hoc MSM analysis with DSM-oriented CBCL sum scores

|  | **Estimate (unstandardized)** | **Robust std. error** | **P value** |
| --- | --- | --- | --- |
| **CBCL anxiety** | | | |
| Age 11-12 chronotype | -0.0003533 | 0.0014924 | 0.815 |
| Age 12-13 chronotype | 0.0003182 | 0.0013154 | 0.811 |
| **CBCL depression** | | | |
| Age 11-12 chronotype | 0.001729 | 0.001548 | 0.278 |
| Age 12-13 chronotype | 0.001056 | 0.001493 | 0.488 |

Table S2. Results of post-hoc MSM analysis stratified by sex: Females

|  | **Estimate (unstandardized)** | **Robust std. error** | **P value** |
| --- | --- | --- | --- |
| **Internalizing** | | | |
| Age 11-12 chronotype | 0.1175 | 0.3626 | 0.74946 |
| Age 12-13 chronotype | 0.2041 | 0.4095 | 0.62396 |
| **Externalizing** | | | |
| Age 11-12 chronotype | -0.03414 | 0.21455 | 0.875268 |
| Age 12-13 chronotype | 0.28883 | 0.21726 | 0.199442 |
| **Prodromal Psychosis** | | | |
| Age 11-12 chronotype | 0.07250 | 0.04076 | 0.0913 |
| Age 12-13 chronotype | 0.03413 | 0.04598 | 0.4670 |

Table S3. Results of post-hoc MSM analysis stratified by sex: Males

|  | **Estimate (unstandardized)** | **Robust std. error** | **P value** |
| --- | --- | --- | --- |
| **Internalizing** | | | |
| Age 11-12 chronotype | -0.1781 | 0.2230 | 0.434 |
| Age 12-13 chronotype | -0.1099 | 0.2342 | 0.644 |
| **Externalizing** | | | |
| Age 11-12 chronotype | -0.2344 | 0.1908 | 0.234 |
| Age 12-13 chronotype | 0.1236 | 0.2274 | 0.593 |
| **Prodromal Psychosis** | | | |
| Age 11-12 chronotype | 0.009442 | 0.011500 | 0.4218 |
| Age 12-13 chronotype | 0.050763 | 0.018133 | 0.0114 |

Table S4. Intraclass Correlation Coefficients^a^

| Internalizing full sample | .006 |
| --- | --- |
| Externalizing full sample | .008 |
| Prodromal Psychosis full sample | .020 |
| CBCL anxiety full sample | .000 |
| CBCL depression full sample | .007 |
| Internalizing females | .000 |
| Externalizing females | .000 |
| Prodromal Psychosis females | .020 |
| Internalizing males | .015 |
| Externalizing males | .016 |
| Prodromal Psychosis males | .031 |

^a^Calculated by fitting a linear mixed-effects model with a fixed overall intercept and a random effect for study site.
